# Parents’ perceptions and acceptability of the human papillomavirus vaccine in rural North-Western Zambia: a descriptive phenomenological study at Kabompo Hospital Affiliated Health Centre

**DOI:** 10.64898/2026.09.14.26362793

**Authors:** Sondashi Bombwe, Sebean Mayimbo, Stella Nankamba

## Abstract

**Background:** Human papillomavirus (HPV) vaccination is the principal primary prevention strategy against cervical cancer, but coverage in Zambia remains uneven and, in Kabompo District, stood at 59% of eligible girls in 2024 against a national and global target of 90%. Because the vaccine is administered to minors, uptake depends on parental consent, which is in turn shaped by parents’ perceptions and acceptability. Existing Zambian evidence is predominantly quantitative and urban. This study explored parents’ perceptions and acceptability of the HPV vaccine in a rural North-Western Zambian setting.

**Methods:** A descriptive phenomenological qualitative study was conducted at Kabompo Hospital Affiliated Health Centre between June and August 2025. We recruited 20 parents and legal guardians of girls aged 9–14 years through maximum-variation purposive sampling. We conducted face-to-face, 20–30-minute in-depth interviews in English or Luvale, guided by a single open-ended core question with theory-informed probes; interviews were audio-recorded and transcribed verbatim. Data were analyzed using Colaizzi’s seven-step method with QDA Miner Lite, and interpreted through Sekhon et al.’s Theoretical Framework of Acceptability. Trustworthiness was addressed through triangulation of transcripts and field notes, member checking, peer debriefing, an audit trail and a reflexive journal.

**Results:** Five themes were generated: knowledge and awareness of the HPV vaccine; perceptions of safety and efficacy; sociocultural and religious influences; the role of health workers; and barriers to access. Awareness that the vaccine prevents cervical cancer was widespread, but knowledge of dosing, mechanism and eligibility was shallow, and several parents first encountered substantive information only at the point of service. Positive appraisals of the vaccine coexisted with persistent fertility-related fears and concerns that the vaccine was “too new”. Preference for herbal remedies, religious framings of divine protection and anxiety that vaccination might licence early sexual activity shaped decisions. Health workers were consistently identified as the most trusted source of information and the decisive influence on consent. Distance, transport costs, fear of injections and non-completion of the schedule limited uptake despite stated willingness.

**Conclusions:** Parental acceptability of the HPV vaccine in this rural setting was high in principle but was attenuated by an intention–behaviour gap driven by misinformation, sociocultural framing and structural burden. Improving coverage will require moving beyond information dissemination to dialogic, nurse-led engagement with parents’ specific concerns, co-designed with traditional and religious leaders, and supported by outreach and dose-tracking systems that reduce the burden of completion.

## Background

Cervical cancer is the fourth most common cancer among women worldwide, with an estimated 660,000 new cases and 350,000 deaths in 2022, and the burden falls disproportionately on low-and middle-income countries [1]. Persistent infection with high-risk human papillomavirus (HPV) types, principally HPV-16 and HPV-18, accounts for approximately 70% of cases [2]. Prophylactic HPV vaccination administered before sexual debut is therefore the single most effective primary prevention tool available, and the World Health Organization’s Global Strategy to Accelerate the Elimination of Cervical Cancer sets a target of vaccinating 90% of girls by the age of 15 years by 2030 [3,4].

Zambia introduced the HPV vaccine in 2013 on a pilot basis in Chongwe, Kafue and Lusaka districts, and rolled it out nationally through the Expanded Programme on Immunization in 2019 [5,6]. Delivery combines a school-based strategy for girls in grade 4 with health centre and community outreach during the annual Child Health Week for out-of-school girls aged 9–14 years [7]. Despite this infrastructure, national coverage has been volatile, falling from 75% in 2020 to 39% in 2021 [12]. In Kabompo District, North-Western Province, only 59% of the 8,436 eligible girls had received the vaccine by 2024 [7], well short of the 90% target. Because the vaccine is given to minors, this shortfall is not principally a supply-side problem: it reflects the decisions of parents and guardians, who hold the power of consent.

Two concepts organize those decisions. Perception refers to how parents understand, interpret and assign meaning to the vaccine, including its purpose, safety, efficacy and necessity. Acceptability, following Sekhon, Cartwright and Francis [8], is a multi-faceted construct reflecting the extent to which people delivering or receiving an intervention consider it appropriate, based on anticipated or experienced cognitive and emotional responses. Their Theoretical Framework of Acceptability specifies seven constructs that are affective attitude, burden, ethicality, intervention coherence, opportunity costs, perceived effectiveness and self-efficacy, that together offer a more textured account of parental decision-making than the binary measure of stated willingness that dominates the HPV literature.

Reported acceptability across sub-Saharan Africa ranges from 59% to 88%, and is strongest where parents receive information directly from health workers and where the vaccine is free [9]. Zambian estimates are similarly favourable in principle: 88% of mothers in Ndola expressed willingness to vaccinate when the vaccine was offered free of charge [10]. Yet stated willingness and actual uptake diverge sharply. Liu et al. [19] reported near-universal hypothetical acceptance in pre-rollout Lusaka, while Nyambe et al. [13] found that post-pilot, only 6.5% of Lusaka parents had vaccinated their daughters. Lubeya et al. [11], applying the Health Belief Model, identified limited knowledge and circulating myths, amplified by the COVID-19 pandemic, as explanations for below-average parental consent. Qualitative work has begun to explain why: Mweemba et al.[14] documented rumours in Lusaka schools that the vaccine contained cervical cancer or caused infertility, and Munjita [15] traced fertility fears, distrust of health authorities and reliance on spiritual and herbal alternatives through Zambian online discourse. Comparable rumours have been reported in Kenya [16], and awareness alone has been shown not to predict willingness in Nigeria [17].

Three gaps remain. First, the Zambian evidence base is overwhelmingly quantitative and cross-sectional, capturing proportions rather than the reasoning parents use to make decisions. Second, it is geographically concentrated in Lusaka and Ndola, urban settings where information, transport and services are more readily available than in rural districts. Third, awareness and knowledge are frequently conflated [18], and acceptability is rarely interrogated with an explicit theoretical framework. No published qualitative study has examined parental perceptions and acceptability of the HPV vaccine in Kabompo District or comparable rural North-Western Zambia, a region with a distinctive ethnic composition and an active calendar of traditional ceremonies. This study addressed these gaps by exploring parents’ awareness and knowledge of the HPV vaccine; their perceptions of its safety, efficacy and benefits; the factors shaping acceptance or rejection; and the role of cultural, religious and social norms in shaping their attitudes.

## Methods

### Study design

A descriptive phenomenological qualitative design, in the Husserlian tradition, was used. Descriptive phenomenology seeks to describe the essence of a phenomenon as it is experienced by those who live it [20,21]. The design suited the research question because perceptions and acceptability are subjective, value-laden experiences that cannot be adequately captured through predefined quantitative variables. The Epoché process bracketed prior assumptions [22], allowing parents’ meanings to emerge in their own terms.

### Study setting

The study was conducted at Kabompo Hospital Affiliated Health Centre, a Ministry of Health primary-level facility in Kabompo Town, the administrative centre of Kabompo District in North-Western Province, Zambia. The district has a population of 70,307, of whom 8,436 are girls aged 9–14 years. The facility is the principal outpatient and community service point affiliated with Kabompo District Hospital, providing maternal and child health, immunization, outpatient, antenatal and postnatal, family planning and HIV services, together with community outreach. The HPV vaccine is offered routinely at the facility and through Child Health Week and school-based outreach. The facility was selected because it is the busiest HPV vaccine delivery point in the district and its catchment includes parents of diverse ethnic, occupational and educational backgrounds, making it an information-rich setting.

### Participants and sampling

Participants were parents and legal guardians of girls aged 9–14 years attending the facility during the data collection period. Eligible participants were aged 18 years or above, had resided in the catchment area for at least six months, could communicate in English or Luvale, and were willing to provide written informed consent for their daughters who could be vaccinated or unvaccinated. Participants were excluded if they were acutely unwell, appeared to be under the influence of alcohol or other substances, had cognitive or communication impairments precluding meaningful engagement with open-ended questions, had participated in pilot interviews, or were health workers, traditional health practitioners or community health volunteers whose professional role could introduce institutional bias into their accounts as ordinary parents.

Maximum-variation purposive sampling was used to include mothers, fathers, and other guardians; parents of vaccinated and unvaccinated daughters; and a range of educational, occupational, and ethnic backgrounds. Twenty participants were recruited. This sample size exceeds the thresholds indicated in the methodological literature, which places code saturation at approximately 9–16 interviews and meaning saturation at 16–24 [24–26]. In practice, we reviewed transcripts and field notes after each interview: code saturation was reached at approximately the fourteenth interview, after which subsequent interviews refined existing themes rather than generating new codes, and meaning saturation was confirmed by the eighteenth. Two further interviews were conducted to confirm saturation before recruitment closed.

### Data collection

Data were collected between June and August 2025. Facility health workers identified parents meeting the inclusion criteria and referred those expressing interest to the researcher, who explained the purpose, procedures, confidentiality measures and voluntary nature of participation, provided an information sheet and obtained written informed consent. Interviews were conducted face-to-face in a private consultation room, lasted 20–30 minutes and were audio-recorded with permission. Consistent with descriptive phenomenology, the semi-structured guide was anchored on a single broad open-ended question inviting participants to describe their experience of the HPV vaccine, supported by sequenced non-leading probes covering awareness, safety and effectiveness, factors influencing acceptance, and cultural, religious and social influences [23]. Interviews were conducted in the participant’s preferred language, English or Luvale. Field notes captured non-verbal expression, emotional tone and contextual detail.

### Data analysis

Recordings were transcribed verbatim, and Luvale interviews were translated into English during transcription while preserving the original meaning. Analysis followed Colaizzi’s seven-step method of descriptive phenomenological analysis [27]: repeated listening and reading for familiarization; extraction of significant statements relating to perceptions and acceptability; formulation of meanings closely tied to participants’ words; clustering of formulated meanings into themes; integration into an exhaustive description; condensation into the fundamental structure of the phenomenon; and member checking of the final findings with selected participants. QDA Miner Lite supported organization, coding, storage and retrieval; interpretation remained the responsibility of the research team. Reflexive notes were maintained throughout and interpretations were discussed regularly with supervisors.

### Trustworthiness and reflexivity

Trustworthiness was addressed through Lincoln and Guba’s four criteria [28]. Credibility was supported by triangulation across verbatim transcripts and field notes, source triangulation across participants of differing ethnic, educational and occupational backgrounds and vaccination status, member checking, peer debriefing with supervisors, and prolonged engagement with the setting. Transferability is supported by thick description of the setting, participants and analytic process. Dependability was supported by an audit trail documenting sampling, interviewing, transcription, coding and theme development, with the interview guide, transcripts, coding frame and analytic memos retained for review. Confirmability was supported by a reflexive journal, the Epoché process before each interview, and the presentation of direct quotations allowing readers to trace interpretations to participants’ words. The lead researcher is a nurse working within the Kabompo health system; this insider position facilitated access and rapport but also carried a risk of social desirability in participants’ accounts, which bracketing, member checking and supervisor debriefing were used to mitigate.

### Ethical considerations

The University of Zambia Biomedical Research Ethics Committee granted ethical clearance (reference 7528-2025), and the National Health Research Authority granted regulatory approval (registration NHRAR-R-3528/22/11/2025). Permission was obtained from Kabompo District Health Office and the facility in-charge. The study was conducted in accordance with the Declaration of Helsinki and the National Health Research Act No. 2 of 2013.

Written informed consent was obtained from all participants; where a participant was unable to read or write, the consent form was read aloud in their preferred language and a thumbprint obtained in the presence of an impartial witness. Participants were assigned codes P1–P20 and no identifying information was recorded on transcripts, recordings or field notes. We stored recordings, transcripts, and field notes on a password-protected, encrypted computer accessible only to the research team; we destroyed the linkage log after completing the analysis. Participants were free to decline any question or to withdraw at any time, and referral to facility counselling services was available had discussion of fertility, culture or religion caused distress.

## Results

### Participant characteristics

Twenty parents and guardians participated. Ages ranged from 26 to 45 years (mean approximately 33 years), with the largest numbers in the 26–30 year (n = 7) and 31–35 year (n = 6) age groups. Thirteen participants had completed secondary education, four tertiary, and three primary. Occupations included market trading (n = 5), business (n = 6), farming (n = 4), teaching (n = 3), civil service (n = 1) and unemployment (n = 1). Ten participants were biological mothers, and ten were other guardians, including aunts, grandmothers, and older sisters, indicating that vaccination decisions for adolescent girls in this setting are frequently made within extended family structures rather than by biological parents alone. Table 1 summarizes participant characteristics.

**Table 1.**
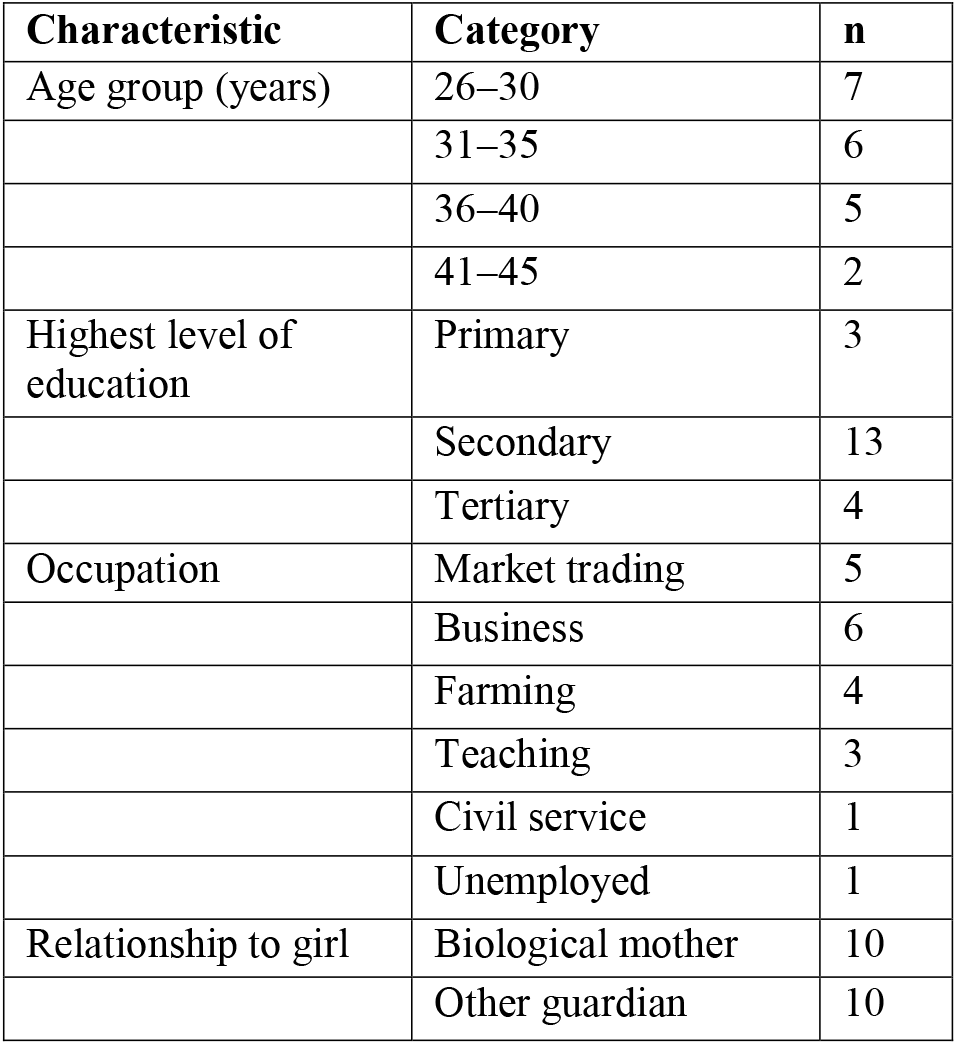
Summary characteristics of participants (n = 20)

| Characteristic | Category | n |
| --- | --- | --- |
| Age group (years) | 26–30 | 7 |
|  | 31–35 | 6 |
|  | 36–40 | 5 |
|  | 41–45 | 2 |
| Highest level of education | Primary | 3 |
|  | Secondary | 13 |
|  | Tertiary | 4 |
| Occupation | Market trading | 5 |
|  | Business | 6 |
|  | Farming | 4 |
|  | Teaching | 3 |
|  | Civil service | 1 |
|  | Unemployed | 1 |
| Relationship to girl | Biological mother | 10 |
|  | Other guardian | 10 |

We generated five themes and thirteen sub-themes from the data (Table 2).

**Table 2.** Themes, sub-themes and representative codes.

| Theme | Sub-theme | Representative codes |
| --- | --- | --- |
| Knowledge and awareness | Basic understanding of the vaccine | Cervical cancer prevention; awareness |
|  | Limited detailed knowledge | Uncertainty; inadequate information |
| Perceptions of safety and efficacy | Positive perceptions | Protection; prevention |
|  | Fear of infertility | Infertility myths; future childbearing |
|  | Fear of side effects | Pain; unknown long-term effects |
| Sociocultural and religious influences | Religious beliefs | Faith healing; divine protection |
|  | Traditional beliefs | Herbal remedies; traditional healers |
|  | Sexual behaviour concerns | Fear of promiscuity |
| Role of health workers | Trusted source of information | Trust; reassurance |
|  | Decision-making support | Guidance; counselling |
| Barriers to access | Distance to facilities | Long travel distance |
|  | Transport challenges | Cost; financial barriers |
|  | Service delivery suggestions | Outreach; door-to-door; radio |

### Theme 1: Knowledge and awareness of the HPV vaccine

Most participants demonstrated basic awareness of the vaccine and correctly identified its purpose as cervical cancer prevention.

> *“It’s for the prevention of cervical cancer.” (P1)*

P10 and P17 gave similar accounts. Participants most frequently attributed their awareness to nurses, schools and Child Health Week campaigns, with health workers described as the point of first introduction to the vaccine.

Awareness, however, did not match depth of understanding. Several participants could not describe the dosing schedule, the mechanism of protection or the relationship between HPV infection and cervical cancer.

> *“I know it is important, but I don’t know how many doses are needed.” (P5)*

For some, substantive engagement with the vaccine occurred only at the point of service, rather than in advance.

> *“I only heard about it when nurses explained.” (P19)*
>
> *“I had no knowledge about it until the nurses explained to me.” (P2, and similarly P20)*

Awareness in this sample was therefore best understood as recognition of the vaccine’s existence and general purpose rather than as a working understanding capable of supporting informed deliberation.

### Theme 2: Perceptions of safety and efficacy

Participants generally appraised the vaccine positively and articulated its protective value.

> *“It is good because it protects our children.” (P13)*
>
> *“It is safe.” (P3)*

This positive baseline coexisted with substantial anxiety. Fear that the vaccine would compromise future fertility was the single most prominent safety concern, and was consistently attributed to information circulating within the community rather than to the health system.

> *“People say girls may fail to have children.” (P14)*
>
> *“People say your child will not be able to conceive.” (P14)*
>
> *“Does it affect fertility?” (P3)*

A second concern was uncertainty about long-term effects, expressed as a sense that the vaccine remained insufficiently proven.

> *“I worry about what it may do later.” (P8)*

Notably, the same participants held positive appraisal of the vaccine’s benefits and serious concern about its safety; recognition of effectiveness did not displace anxiety, and these concerns were reported to have delayed uptake.

### Theme 3: Sociocultural and religious influences

Religious conviction shaped decisions, most often through the belief that protection from disease belongs to the spiritual rather than the medical domain.

> *“We believe God protects us from diseases.” (P12)*
>
> *“Some religious groups refuse to accept because they believe that there is no such disease.” (P10)*

Traditional explanatory frameworks operated alongside biomedical ones, with some participants reporting a community preference for herbal treatment over injection.

> *“Some people just believe in herbal remedies instead of injections.” (P5, and similarly P8 and P11)*

A further concern was moral rather than biomedical: that vaccinating young girls against a sexually transmitted infection might be read as licensing early sexual activity.

> *“It may make girls become promiscuous.” (P7)*

These influences were not uniform or fixed. Several participants had reconciled religious conviction with the decision to vaccinate, typically citing the recommendation of a trusted health worker or a respected community figure as the factor that resolved the tension.

### Theme 4: The role of health workers

Health workers were identified as the most trusted source of vaccine information and, for many participants, the decisive influence on consent.

> *“We trust what nurses tell us.” (P2)*

Participants linked their acceptance directly to the quality of explanation received, emphasizing clarity and the opportunity to have questions answered rather than the mere fact of a recommendation.

> *“If nurses explain properly, we accept.” (P6)*

Trust in health workers therefore functioned as the principal mechanism through which community-level rumor was countered, and participants who had encountered fertility-related claims described nurse explanation as what enabled them to proceed.

### Theme 5: Barriers to access

Although the vaccine was described as generally available through schools and outreach, participants identified practical constraints that limited uptake and, in particular, completion.

> *“The clinic is very far.” (P9)*
>
> *“We fail because transport money is difficult.” (P11)*

Fear of injections among the girls themselves, and difficulty returning for a subsequent dose, were also reported. Several participants who expressed willingness to vaccinate described daughters who had received an initial dose but had not returned, or who remained unvaccinated despite parental consent in principle.

Participants offered concrete suggestions for improving delivery, centering on bringing services closer to households and strengthening community-level information.

> *“They should bring vaccines door-to-door.” (P3)*

Others recommended intensified radio sensitisation in local languages and more frequent community outreach.

## Discussion

This study explored parents’ perceptions and acceptability of the HPV vaccine in a rural North-Western Zambian district where coverage remains well below target. The central finding is that acceptability was high in principle yet incompletely realised in practice: parents endorsed the vaccine, trusted the health workers who offered it, and nonetheless hesitated, delayed or failed to complete the schedule. Understanding that gap requires attention to knowledge, to rumour, to sociocultural framing and to structural burden simultaneously.

Awareness of the vaccine was widespread, indicating that the communication channels established by the Ministry of Health and Kabompo District Health Office are reaching the community, consistent with improvements documented across sub-Saharan Africa since national introduction [9,17]. Depth of knowledge, however, was uneven, mirroring findings that parental knowledge gaps persist more than a decade after introduction [18] and that depth of understanding tracks educational background more closely than awareness alone [29]. The observation that several participants first learned about the vaccine at the point of service is consequential: it contrasts with high-income settings where pre-vaccination paediatric consultation is routine [18], and it means that parents in this setting are frequently asked to consent without prior opportunity for deliberation. The wider implication is conceptual as well as practical. Awareness and knowledge are distinct constructs, frequently conflated in the Zambian literature [36], and Anyaka et al. [17] have shown that awareness does not predict willingness. Increasing the volume of awareness messaging is therefore unlikely to be sufficient; what is required is communication that builds genuine understanding of how HPV causes cervical cancer, why early vaccination matters and what the established safety record is.

Fertility-related fear was the dominant safety concern, despite the absence of any biomedical link between HPV vaccination and infertility [4]. This is among the most consistently reported findings in the African literature: Vermandere [16] documented rumors in Kenya that the vaccine was a covert form of population control; Mweemba et al. [14] found fertility rumours to be among the most influential drivers of hesitancy in Lusaka, communicated through informal networks rather than formal channels; and Munjita [15] identified fertility concerns and conspiracy theories as dominant themes in Zambian online discourse amplified by COVID-19-era distrust. The present findings confirm that this pattern extends into rural North-Western Zambia, where digital penetration is lower and information travels through different channels. Concern that the vaccine was “too new” echoes evidence from high-income settings, where safety concerns are the leading barrier reported by clinicians [30] and where parents systematically underestimate the vaccine’s response efficacy relative to objective benchmarks [31]. The persistence of this concern more than a decade after introduction in Zambia suggests that long-term safety information is either not reaching parents or is not being communicated in a form they find meaningful.

A methodologically important observation is that perceived safety and perceived effectiveness operated as separable constructs in participants’ reasoning, as Myhre et al. [31] demonstrated quantitatively. Parents who could articulate the vaccine’s benefit in preventing cervical cancer simultaneously held serious safety concerns. This directly undermines the information-deficit model that implicitly underpins much health education practice. Interventions must engage the specific anxiety parents already hold — above all, fertility — rather than simply supplying more facts about benefit, and must do so through sources the community already trusts, without dismissing the underlying concern as ignorance.

Stated acceptability was high, consistent with the 59–88% range reported across sub-Saharan Africa [9], the 88% maternal willingness reported in Ndola when the vaccine was free [10], and evidence that institutional endorsement raises acceptability substantially [32]. Yet the intention–behaviour gap was clearly visible, paralleling the contrast between near-universal hypothetical acceptance in pre-rollout Lusaka [19] and 6.5% actual uptake following the pilot [13], and the consent–follow-through gap identified by Lubeya et al. [11]. Read through Sekhon et al.’s framework [8], the present findings suggest several constructs operating at once: affective attitude was positive; intervention coherence was uneven; perceived effectiveness was high in principle but vulnerable to rumour; ethicality was contested where the vaccine was associated with adolescent sexuality; and burden such as distance, transport cost and the requirement to return was substantial. Programmes that measure willingness alone, without addressing coherence, ethicality and burden, are unlikely to close this gap.

Sociocultural and religious influences operated largely below the threshold of formal health system communication. Preference for herbal remedies reflects the coexistence of biomedical and traditional explanatory frameworks widely documented in sub-Saharan Africa, and is consistent with reports that a substantial proportion of parents in rural Zambia attribute cervical cancer to witchcraft rather than to HPV infection, rendering biomedical prevention irrelevant within their own explanatory logic [6]. Religious framings of divine protection align with Vermandere’s Kenyan findings [16] and with the Lusaka case study of Mweemba et al. [14]. Concern about sexual disinhibition has been identified as one of the most consistently reported barriers in systematic reviews of parental attitudes [33,34]. Critically, however, these influences were dynamic rather than fixed: participants who had reconciled religious conviction with vaccination typically cited a trusted health worker or community leader as the resolving factor. This supports the argument that cultural and religious factors are processes open to respectful dialogue rather than static barriers [34], and implies that effective interventions in Kabompo must be codeveloped with traditional leaders, religious authorities and respected community members rather than designed in their absence.

The determinative role of health workers was the clearest actionable finding. Trust in nurses was near-universal, and participants linked acceptance explicitly to the quality of explanation received. This aligns with evidence that strong, unambiguous provider recommendation is the single most important predictor of HPV vaccine uptake [30], that health worker advice significantly increases parental consent in Zambia [11], and that health workers can either reinforce or counteract community rumour [14]. Availability of the vaccine is therefore necessary but not sufficient: health workers require current evidence on safety and efficacy, communication tools in local languages, and above all the protected time to engage in genuine dialogue with parents holding fertility-related or religious concerns.

Finally, access barriers — distance, transport cost, fear of injections and non-completion — were consistent with the logistical and infrastructural constraints documented in Zambian programme delivery evaluations [6]. Participants’ own suggestions of door-to-door delivery, intensified community sensitisation and expanded radio education point towards a more active service delivery model that reduces travel burden while improving information flow.

### Strengths and limitations

To the authors’ knowledge, this is the first qualitative study of parental perceptions and acceptability of the HPV vaccine in Kabompo District and one of very few in rural North-Western Zambia, addressing a clear geographical gap. The explicit application of Sekhon et al.’s Theoretical Framework of Acceptability [8] addresses a conceptual gap in the Zambian literature, where acceptability has commonly been operationalized loosely as willingness to vaccinate. The descriptive phenomenological approach, anchored on a single open-ended core question, surfaced concerns such as fertility fears, religious framings of protection, and moral anxiety about adolescent sexuality that structured survey instruments would likely have flattened. Maximum-variation sampling and systematic attention to trustworthiness, including triangulation, member checking, peer debriefing, an audit trail and reflexive journaling, support confidence in the findings.

Several limitations should be borne in mind. The study was conducted at a single facility, and parents in remote zones who attend other health posts or who do not regularly access services were under-represented; findings reflect the experiences of facility attenders rather than the district as a whole. The sample was predominantly female, reflecting the pattern in which mothers and female guardians attend health facilities, so the perspectives of fathers and male guardians are proportionately fewer. The lead researcher’s position as a nurse within the local health system may have shaped question framing and interpretation, and some participants may have moderated criticism of the health system in the presence of a known health worker. Interviews conducted in Luvale were translated during transcription, and some idiomatic nuance may have been lost despite member checking and supervisor review. Finally, the descriptive phenomenological design provides depth rather than prevalence: the findings should be read as a context-specific account of meaning-making, not as a generalisable estimate of the distribution of attitudes.

## Conclusions

Parental acceptability of the HPV vaccine at Kabompo Hospital Affiliated Health Centre was high in principle but incompletely realised in practice. Awareness was widespread while knowledge was shallow; positive appraisals of the vaccine coexisted with persistent fertility-related fears; sociocultural and religious frameworks shaped decisions through channels the health system does not routinely reach; health workers were the most trusted and decisive influence; and distance, cost and the burden of completion attenuated stated willingness. Improving coverage in this setting will require moving beyond information dissemination towards dialogic, nurse-led engagement with the specific concerns parents already hold, supported by refresher training in HPV communication, protected counselling time, dose reminder and tracking systems, and outreach that reduces the burden of return visits. Community sensitisation should be co-designed with traditional leaders, religious authorities and parents who have vaccinated their daughters, and should engage fertility myths openly and respectfully rather than dismissing them. Further qualitative work in other rural districts, longitudinal mixed-methods research tracking decision-making across the full schedule, intervention studies testing nurse-led dialogic communication, and research on the role of male parents and grandparents are all warranted.

## Data Availability

The data underlying this study cannot be shared publicly due to ethical restrictions and concerns regarding participant confidentiality. The dataset consists of transcripts from in-depth qualitative interviews that contain sensitive personal beliefs and potentially identifying contextual information. Public sharing of these transcripts would compromise the privacy of the parents and guardians who participated, violating the conditions of the informed consent and the ethical approval granted by the University of Zambia Biomedical Research Ethics Committee (UNZABREC). De-identified, representative excerpts of the data are available within the manuscript. Researchers who meet the criteria for access to confidential qualitative data may submit data access requests to the University of Zambia Biomedical Research Ethics Committee (UNZABREC) at, or by mail at Ridgeway Campus, P.O. Box 50110, Lusaka, Zambia.

## Declarations

### Ethics approval and consent to participate

The University of Zambia Biomedical Research Ethics Committee (reference 7528-2025) granted ethical clearance, and the National Health Research Authority of Zambia (registration NHRAR-R-3528/22/11/2025) granted regulatory approval. Permission was obtained from Kabompo District Health Office and Kabompo Hospital Affiliated Health Centre. All participants provided written informed consent; participants unable to read or write gave a thumbprint after the consent form was read aloud in their preferred language in the presence of an impartial witness. The study was conducted in accordance with the Declaration of Helsinki and Zambia’s National Health Research Act No. 2 of 2013.

### Consent for publication

Not applicable. No individually identifying information is reported; participants are identified only by anonymised codes.

### Availability of data and materials

The anonymised transcripts generated and analysed during the current study are not publicly available in order to protect participant confidentiality in a small rural community, but are available from the corresponding author on reasonable request and subject to approval by the University of Zambia Biomedical Research Ethics Committee. The interview guide is available from the corresponding author on request.

### Competing interests

The authors declare no competing interests.

### Funding

This study was self-funded by the lead author. No external funding was received for the design of the study, data collection, analysis, interpretation of data, or writing of the manuscript.

### Authors’ contributions

SB conceived and designed the study, collected and analysed the data, and drafted the manuscript. SM and SN supervised the study, contributed to the design and analytic framework, participated in peer debriefing and review of emerging themes, and critically revised the manuscript. All authors read and approved the final manuscript.

## Acknowledgements

The authors thank the Kabompo District Health Office for granting permission to conduct the study, the staff of Kabompo Hospital Affiliated Health Centre for their assistance with participant identification, and above all the parents and guardians who gave their time and shared their experiences.

## Notes

### Competing Interest Statement

The authors have declared no competing interest.

### Author Declarations

University of Zambia Biomedical Research Ethics Committee gave approval for this work

